# Combined Gastric Alimetry® and gastric emptying scintigraphy testing increases clinician certainty in the diagnosis and management of suspected gastroparesis

**DOI:** 10.1101/2025.05.06.25327029

**Authors:** Ryan Abraham, Daphne Foong, Vincent Ho

## Abstract

**Background and Aim:** Gastric emptying scintigraphy (GES) is the reference standard test for diagnosing gastroparesis. Body surface gastric mapping (BSGM) via Gastric Alimetry® is a new test of gastric function that combines non-invasive assessment of gastric electrophysiology and validated symptom profiling. This randomized, prospective pilot study evaluated the impact of GES vs BSGM test results on clinical decision-making.

**Methods:** Patients with chronic gastroduodenal symptoms from a tertiary center referred for GES were recruited. Subjects separately underwent baseline assessment with GES and BSGM testing. Two motility-specialists were first asked to devise a management plan after reviewing a test result (GES or BSGM, in random order). They were then asked to repeat the management plan after reviewing the other test result (BSGM or GES). Clinician-perceived certainty measures were assessed.

**Results:** Sixteen patients, 13 (81%) female, median age 30 years, median BMI 22.5 kg/m^2^, were recruited. At baseline, a diagnosis was established in 2/16 (12.5%) and increased to 8/16 (50%) with both tests. Abnormal test results were found in 11 patients. In patients with normal results, BSGM symptom profiling phenotyped 5 additional patients. All patients received an intervention following the first unblinding, with subsequent management changes made in 75% (BSGM) and 62.5% (GES) of patients. The combined GES and BSGM results significantly increased diagnostic and management certainty (p<0.05), with both tests having similar influence on management (p>0.05).

**Conclusion:** The combined GES and BSGM test results significantly enhanced diagnostic and management confidence in patients with suspected gastroparesis within a tertiary center.

## Introduction

Gastroparesis is a chronic and debilitating gastric disease currently defined as persistent upper gastrointestinal symptoms and delayed gastric emptying with no mechanical gastric outlet obstruction ^1^. The condition is associated with poor quality of life, psychological distress, frequent hospitalizations, and consequent high healthcare utilization and associated costs ^2–6^. Despite the significant clinical burden, current diagnostic and management approaches remain suboptimal, with only 30% of patients experiencing symptom improvement following standard care ^7^. This challenge in effectively managing chronic symptoms is further exacerbated by the persistently high economic burden placed on these patients, which remains significantly elevated compared to controls even years after initial diagnosis and treatment ^5^.

Gastric emptying scintigraphy (GES) remains the reference standard for diagnosing gastroparesis ^8^. However, its clinical utility has been increasingly questioned due to its variability over time and poor correlation with symptoms ^9,10^. GES provides a detailed measure of gastric emptying rate from the stomach, but provides limited insight into the underlying pathophysiological mechanisms, potentially explaining why the effectiveness of current management strategies based on emptying alone often yield poor results ^1,5,7,9,11^.

Body surface gastric mapping (BSGM) using Gastric Alimetry® (Alimetry Ltd., New Zealand) is a new test of gastric function based on non-invasively assessing gastric motility using simultaneous high-resolution electrogastrography and validated symptom profiling ^12–14^. Previous research has demonstrated BSGM’s clinical utility in the assessment of gastric function through patient phenotyping in various gastroduodenal disorders, including patients with nausea and vomiting disorders, diabetes, delayed gastric emptying, and post-gastric surgery conditions ^15–20^. Notably, within these populations the detection of gastric motility abnormality rates were higher using Gastric Alimetry compared to GES alone (33% vs 23%), suggesting the potential of BSGM to enhance diagnostic precision and guide more targeted treatment ^19^. The clinical application of BSGM and its impact on management have been previously described in various studies; the results showed that BSGM-guided management led to a significant reduction in healthcare utilization and cost ^21–23^.

While both GES and BSGM provide information about gastric function, it remains unclear how these two diagnostic approaches differentially influence clinical management decisions. Previous studies have investigated the impact of various motility tests on clinical management. These studies assessed the change in management, diagnosis or investigations based on antroduodenal manometry results ^24^ and on simultaneous GES and wireless motility capsule investigations ^25^. Based on these designs, the present study aimed to evaluate the impact of GES vs BSGM test results on clinical decision-making through an exploratory, two arm, randomized, prospective trial in patients with suspected gastroparesis undergoing GES and BSGM testing. We hypothesized that incorporating BSGM testing would significantly alter clinical management decisions, and increase clinician diagnostic and therapeutic certainty compared to GES alone.

## Methods

### Patient population

Ethics approval was obtained from Western Sydney University Human Research Ethics Committee (H15874), and the study has been performed in accordance with the Declaration of Helsinki (2013). All patients provided informed consent. Patients aged ≥ 18 years old and referred to GES for evaluation of their chronic gastroduodenal symptoms were recruited from a tertiary center. Upper gastrointestinal endoscopy was performed to exclude organic disorders. Exclusion criteria included pregnancy, breastfeeding, inability to undergo BSGM testing due to history of adhesive allergies or damaged epigastric skin, or having previously undergone a BSGM test.

### Test methodologies

Each subject separately underwent baseline assessment with GES and BSGM testing, as per standard protocol ^8,15^. All patients were fasted for at least 6 hours, and any GI medications were withheld for at least 48 hours.

GES consisted of a standard ∼255 kCal low-fat egg meal radiolabeled with ^99m^Tc followed by imaging using a gamma camera over 4 hrs. GES results were interpreted using standardized criteria ^8^: delayed if >10% retention at 4 hrs, or rapid if <30% retention at 1 hr.

As previously described, BSGM testing encompassed a 30-minute fasting recording followed by a standard 480 kCal test meal and 4 hr postprandial recording ^15^. Throughout the study, patients used the Gastric Alimetry app to rate their upper gastroduodenal symptoms every 15 minutes on a 0 - 10 Likert scale ^14^ and to complete 10 close-ended questions on an Alimetry Gut-Brain Wellbeing (AGBW) Survey at the start of the test, which assesses mental wellbeing including depression, anxiety and stress ^26^. BSGM spectral metrics were compared to validated normative reference intervals ^27^: Gastric Alimetry Rhythm Index (GA- RI), BMI-adjusted amplitude, principal gastric frequency (PGF) and Fed:Fasted Amplitude Ratio. BSGM symptom profiles were categorized using a standardized classification framework based on previous publications ^19,28^.

Symptoms, quality of life and health psychology were also evaluated using validated questionnaires: Rome IV criteria ^29^, PAGI-SYM ^30^, GCSI ^31^, PAGI-QOL ^32^, PHQ-9 ^33^, GAD-7 ^34^, and PSS-4 ^35^.

### Management plan

The method was similar to those used in previous studies ^24,25^. Prior to testing, an initial management plan was in place based on the patient’s presenting symptoms and clinical history. Following testing and before each patient’s follow up appointment, two motility-specialist gastroenterologists reviewed and interpreted the GES and BSGM test results. BSGM report interpretation was performed by an independent experienced reviewer following established test interpretation guidelines ^36^.

Two management plans were completed using standardized forms. After reviewing the patient’s clinical history and initial management plan, clinicians were first provided a test result (GES or BSGM, in random order) and asked to devise a management plan for the patient (Plan 1), including recommendations for medications, diet, surgery referral, other intervention, or additional testing. They were then given the other test result (BSGM or GES) and asked to repeat the management plan with the new combined information (Plan 2). The method flow chart is shown in Supplementary Figure 1.

### Clinician-perceived utility

Clinicians rated diagnostic and management plan certainty using a Likert Scale (0-10; 0=uncertain, 10=certain) firstly after reviewing the initial allocated test (GES or BSGM) and formulating Plan 1, and again after reviewing the second test and formulating Plan 2. At the conclusion, clinicians were asked to rate the level of contribution of each test result in terms of their insights to the clinical diagnosis and usefulness of information for devising the plan from 0 (least) to 5 (most), and whether the test results aligned (yes/no). This similar approach to capturing clinician-perceived utility has been previously used in studies examining the clinical utility of diagnostic testing ^37–39^.

### Data analysis

The primary endpoint was the change in clinical management decisions from Plan 1 to Plan 2 due to the combined test results. This methodology was based on approaches used in previous studies ^24,25^. In Plan 1, a management change was defined as any addition, removal or modification within a category (e.g., switching from one prokinetic to another) compared to the initial management plan. Medication changes were categorized into specific categories: prokinetics, antiemetics, neuromodulators, proton pump inhibitors, laxatives, and other. Diet recommendations were classified as gastroparesis diet, FODMAP diet, referral to dietician or other. Psychology-based changes were categorized as referral to psychology or gut hypnotherapy. Any referrals to surgery and additional testing were recorded. All other changes were referred to as other interventions.

In Plan 2, a change due to the combined test results was defined as any addition, removal or modification within the same category compared to Plan 1. A change in diagnosis was defined as either an addition of a diagnosis or a shift to a different one (confirmation of an existing diagnosis was not considered a change).

Similar to the approach used by Hasler et al. (2019) ^25^, we also analyzed how specific test findings related to management decisions by examining the relationship between the test result subgroups (normal/abnormal) and changes in management decisions.

### Statistical methods

Data are presented as median with interquartile range (IQR). Continuous data were compared using paired t test. Categorical data were assessed using Fisher’s exact test; p-value < 0.05 was considered statistically significant.

## Results

### Study population

Sixteen consecutive patients completed the study: 13 females (81%), 3 males (19%); median age 30 years (IQR 22 - 59); median BMI 22.45 kg/m^2^ (IQR 18.43 - 27.75). Three patients (19%) had diabetes. There were no differences in any clinical variables between the two randomized arms (GES then BSGM vs BSGM then GES; p>0.05; Table 1). Prior to testing, 94% of patients met Rome IV criteria for both chronic nausea and vomiting syndrome (CNVS) and functional dyspepsia (FD); one patient met criteria for CNVS only. Patient clinical variables is shown in Table 1.

**Table 1:** Baseline patient demographics. P-values represent comparisons between GES then BSGM and BSGM then GES.

| Variable | Total | GES then BSGM | BSGM then GES | P-value |
| --- | --- | --- | --- | --- |
| Age, median (IQR) | 30.00 (22.25-59.00) | 49.00 (31.50-61.00) | 23.00 (18.00-32.25) | 0.12 |
| BMI, median (IQR) | 22.45 (18.43-27.75) | 24.65 (21.50-27.15) | 19.15 (16.20-28.20) | 0.50 |
| Gender, female, n (%) | 13 (81.2) | 7 (87.5) | 6 (75.0) | 1.00 |
| Gender, male, n (%) | 3 (18.8) | 1 (12.5) | 2 (25) | 1.00 |
| Diabetes, n (%) | 3 (18.8) | 2 (25.0) | 1 (12.5) | 1.00 |
| PAGI-SYM score, median (IQR) | 1.94 (1.10-2.23) | 1.94 (1.01-2.22) | 2.03 (1.28-2.39) | 0.50 |
| GCSI score, median (IQR) | 2.31 (1.49-2.35) | 1.89 (0.94-2.32) | 2.32 (1.74-2.74) | 0.13 |
| PAGI-QOL score, median (IQR) | 3.49 (3.25-3.69) | 3.51 (3.45-3.79) | 3.48 (3.06-3.65) | 0.91 |
| PSS-4 score, median (IQR) | 7.00 (4.50-8.50) | 8.00 (7.50-8.50) | 5.00 (3.25-6.00) | 0.42 |
| GAD-7 score, median (IQR) | 6.00 (2.50-7.50) | 6.00 (5.00-7.00) | 5.00 (2.00-8.00) | 0.96 |
| PHQ-9 score, median (IQR) | 7.00 (5.00-13.50) | 12.00 (5.50-14.50) | 6.50 (5.00-10.00) | 0.56 |

### Test results

Combined GES+BSGM profiling enhanced patient classification compared to single-test approaches (Figure 1A). While GES alone provided definitive classification in 2/8 patients (25%), the combined GES+BSGM approach classified 8/8 patients (100%, p=0.007). In contrast, there was no difference in the classification between BSGM alone vs combined BSGM+GES (8/8 patients; p>0.05).

**Figure 1:**
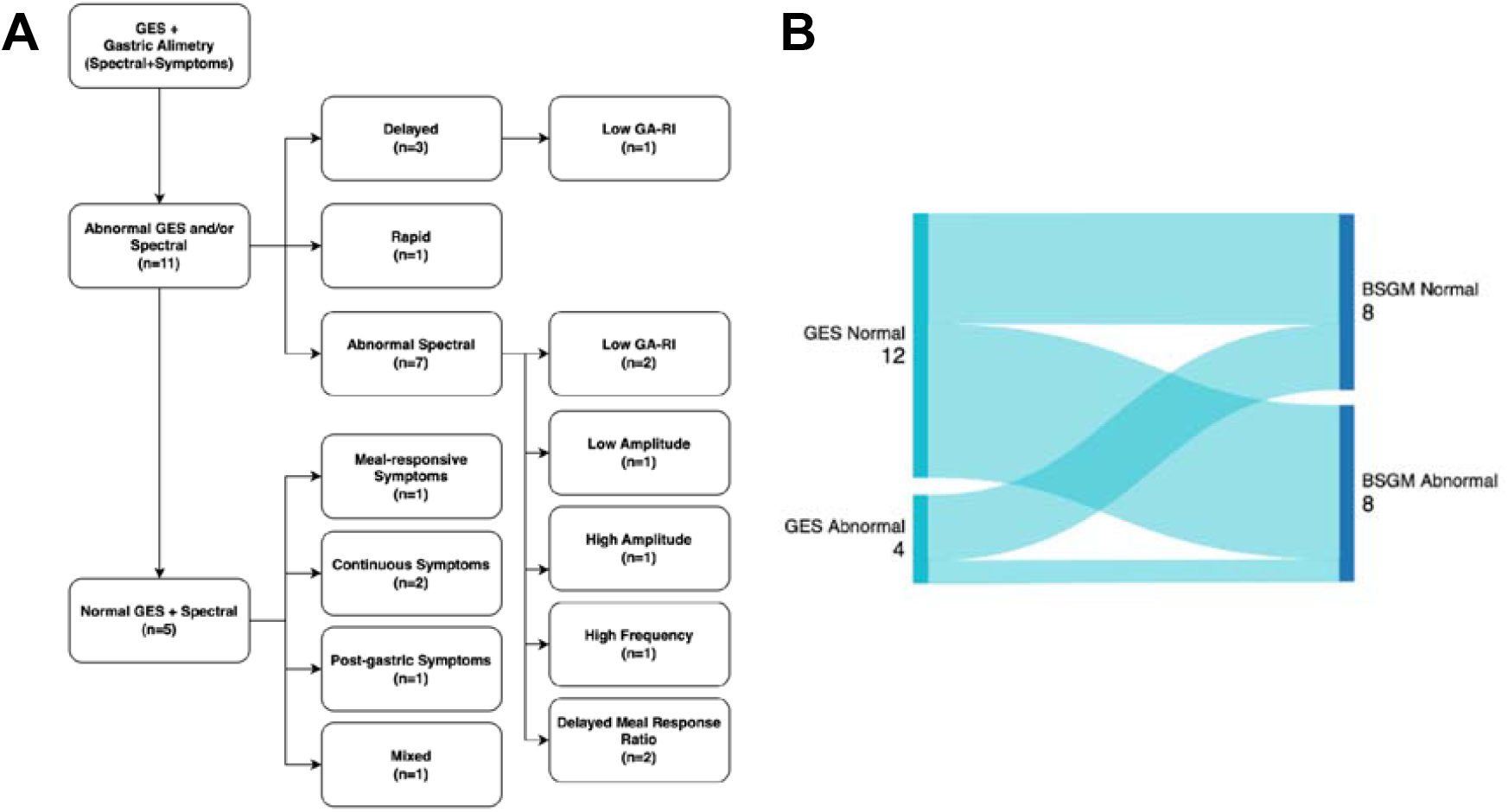
Summary of overall test results. A) Combined GES+BSGM profiling led to increased classification of patients. B) Sankey plot showing little overlap between GES and BSGM test results.

Among the 11 patients with abnormal test results (69%), the distribution of abnormalities demonstrated minimal overlap between the two testing modalities (Figure 1B): 3 had delayed or rapid GES, 7 had abnormal spectral analysis on BSGM only, and 1 patient had both a delayed GES and abnormal spectral analysis (Figure 1B). BSGM spectral abnormalities included low GA-RI (n=3), low amplitude (n=1), high amplitude (n=1), high frequency (n=1) and delayed meal response ratio (n=2).

Of the remaining 5 patients (31%) with normal GES and normal spectral analysis on BSGM, BSGM symptom profiling provided additional phenotypic information based on standard classification criteria ^19,28^: meal-responsive (n=1), continuous (n=2), post-gastric (n=1) and mixed patterns (n=1).

### Change in clinical diagnosis

At baseline, use of only the patient’s clinical history provided a single diagnosis in 2/16 cases (12.5%; Figure 2). After unblinding one test result (either GES or BSGM), this increased to 3/16 cases (18.8%). Following review of both test results, clinicians were able to establish a single definitive diagnosis in 8/16 cases (50%, p=0.054 compared to baseline).

**Figure 2:**
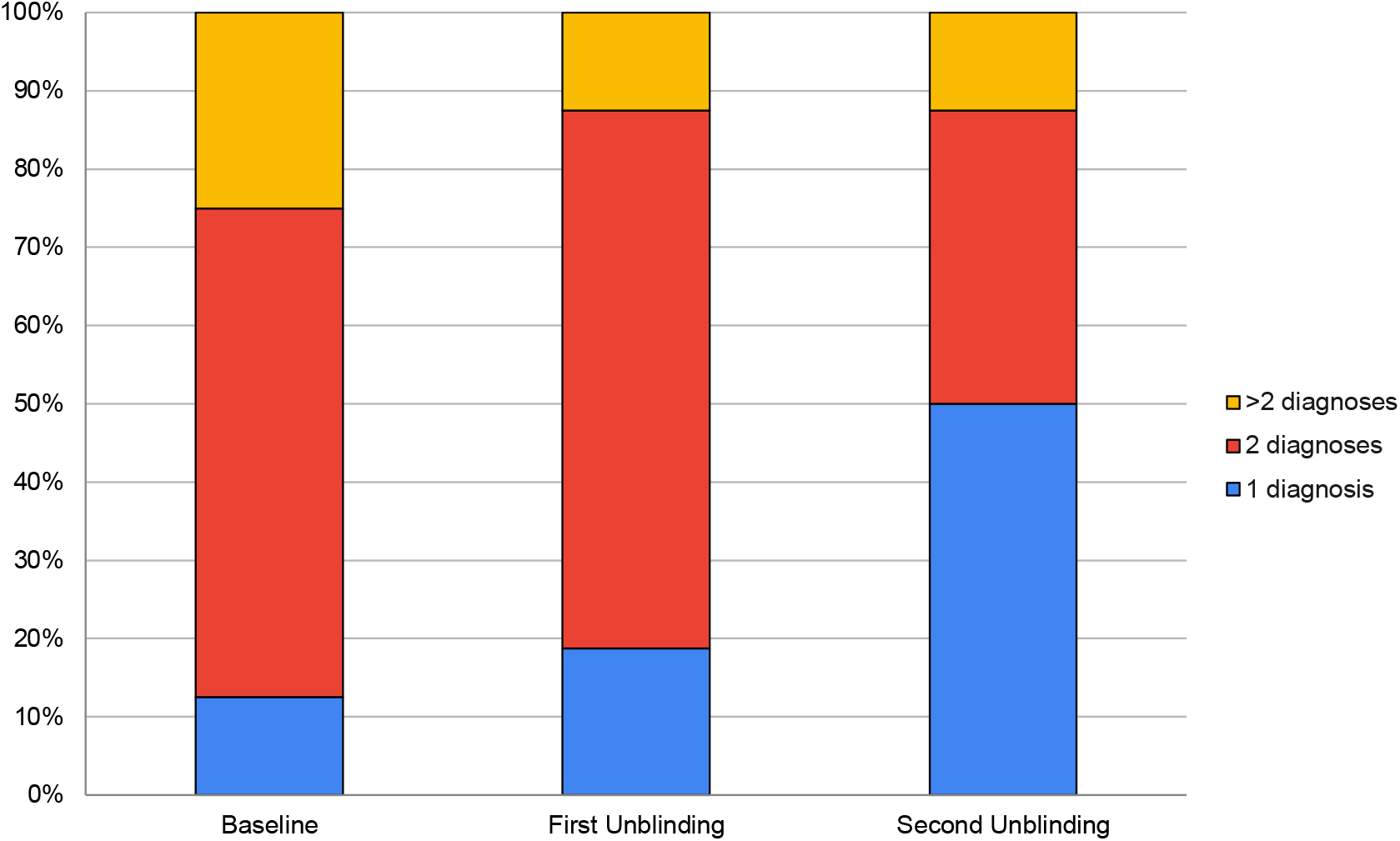
Change in clinical diagnosis after each unblinding. Numbers are the percentage of total patients.

Final diagnoses included functional dyspepsia (n=4), gastroparesis (n=1), cyclic vomiting syndrome (n=1), and non-gastroduodenal disorders (n=2: acute illness and median arcuate ligament syndrome). The remaining patients were diagnosed with a combination of functional dyspepsia, gastroparesis, Rome-IV gastroduodenal disorder, other gut motility disorder and/or eating disorder.

### Change in clinical management decisions

Following review of the first test result (either GES or BSGM), clinicians recommended therapeutic interventions for all patients (Table 2). Initial management changes were similar between the test results, including medication changes in 75% of patients, diet modifications in >50% of patients, and psychology-based interventions in 12.5% of patients (GES vs BSGM; p>0.05).

**Table 2:** Changes in clinical management due to the test results. Numbers are the percentage of total patients.

|  | GES then BSGM (n=8) |  | BSGM then GES (n=8) |  |
| --- | --- | --- | --- | --- |
|  | GES | GES+BSGM | BSGM | BSGM+GES |
| **Overall Change** | 100 | 75 | 100 | 62.5 |
| **Medication Change** | 75 | 37.5 | 75 | 37.5 |
| **Diet Change** | 62.5 | 12.5 | 50 | 25 |
| **Psychology-based interventions** | 12.5 | 25 | 12.5 | 12.5 |
| **Surgery referral** | 0 | 0 | 12.5 | 0 |
| **Other intervention change** | 12.5 | 12.5 | 0 | 0 |
| **Additional testing change** | 0 | 12.5 | 0 | 0 |

The additional unblinding of the second test result led to substantial management changes in both arms of the study: 75% of patients in the ‘GES then BSGM’ arm and 62.5% in the ‘BSGM then GES’ arm received further management modifications. These changes included medication adjustments (37.5% in both arms), dietary modifications (12.5% in ‘GES then BSGM’ arm; 25% in ‘BSGM then GES’ arm), and psychology-based interventions (25% in ‘GES then BSGM’ arm; 12.5% in ‘BSGM then GES’ arm).

Common medication recommendations included prokinetics such as Domperidone and Metoclopramide, neuromodulators such as Amitriptyline, and other medications such as Iberogast (STW5). Notable non-pharmacological management decisions following the combined test results included: referral to upper gastrointestinal surgery (n=1), discontinuation of Semaglutide with endocrinology referral (n=1), geriatric referral (n=1), and referral for manometry and 24-hour pH testing (n=1).

### Change in management decisions in relation to the test results

Six patients were recommended prokinetics. For three patients, prokinetics were recommended following unblinding of a delayed emptying result, with two of the three patients also recommended a gastroparesis diet. These recommendations remained the same after unblinding BSGM regardless of the test results. The remaining three patients with prokinetic recommendations had normal GES test results, but two of three patients had an abnormal BSGM: borderline high amplitude (n=1) and delayed meal response ratio (n=1). All three patients who were recommended a neuromodulator had normal gastric emptying. Two of three patients were prescribed a neuromodulator due to the continuous symptom profile or AGBW scores on BSGM. Five patients were recommended the herbal tincture Iberogast, mostly as a result of their meal-responsive or sensorimotor symptom profile. The individual changes in clinical diagnosis and management decisions after each unblinding are shown in Supplementary Table 1.

### Clinician perceived certainty and utility

Combined GES+BSGM test results significantly enhanced clinician certainty compared to single-test results (Figure 3). Diagnostic certainty increased from a median of 6.5 (IQR 5.8-8.0) with GES alone to 8.0 (IQR 7.8-9.0) with GES+BSGM (p=0.0008), and from 5.0 (IQR 3.0-7.3) with BSGM alone to 8.0 (IQR 7.8-9.0) with BSGM+GES (p=0.008).

**Figure 3:**
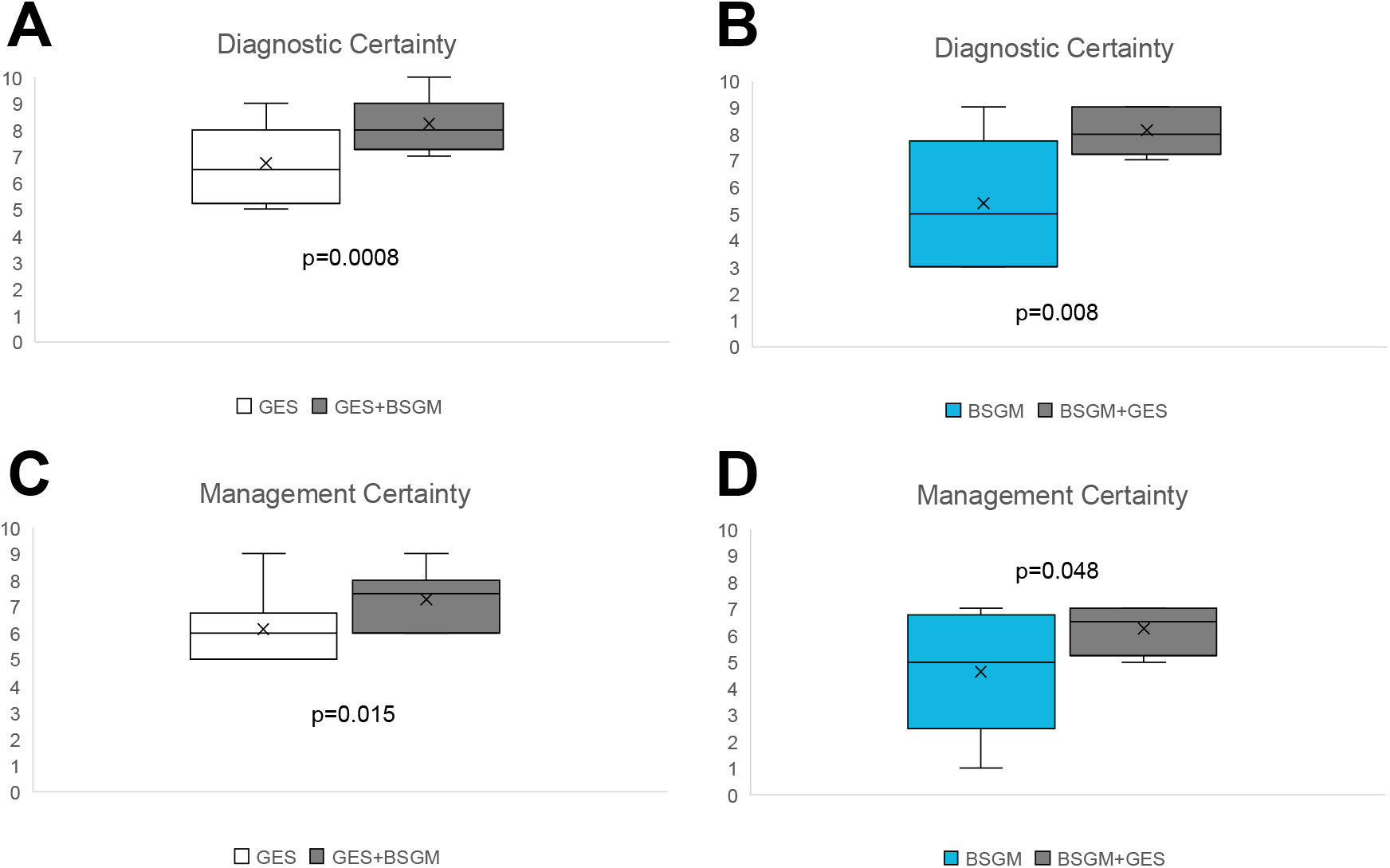
Diagnostic (A, B) and management (C, D) certainty of GES vs GES+BSGM and BSGM vs BSGM+GES. Statistical comparisons were analyzed using paired t test.

Similarly, management certainty improved from a median of 6.0 (IQR 5.0-6.3) with GES alone to 7.5 (IQR 6.0-8.0) with GES+BSGM (p=0.015), and from 5.0 (IQR 3.5-6.3) with BSGM alone to 6.5 (IQR 5.8-7.0) with BSGM+GES (p=0.048).

There were no differences in diagnostic and management certainty between GES and BSGM when the tests were considered in isolation (p>0.05). Furthermore, clinicians perceived alignment between test results in 81% of cases. Both tests were deemed equally contributory to diagnostic and management decisions (p>0.05; Figure 4).

**Figure 4:**
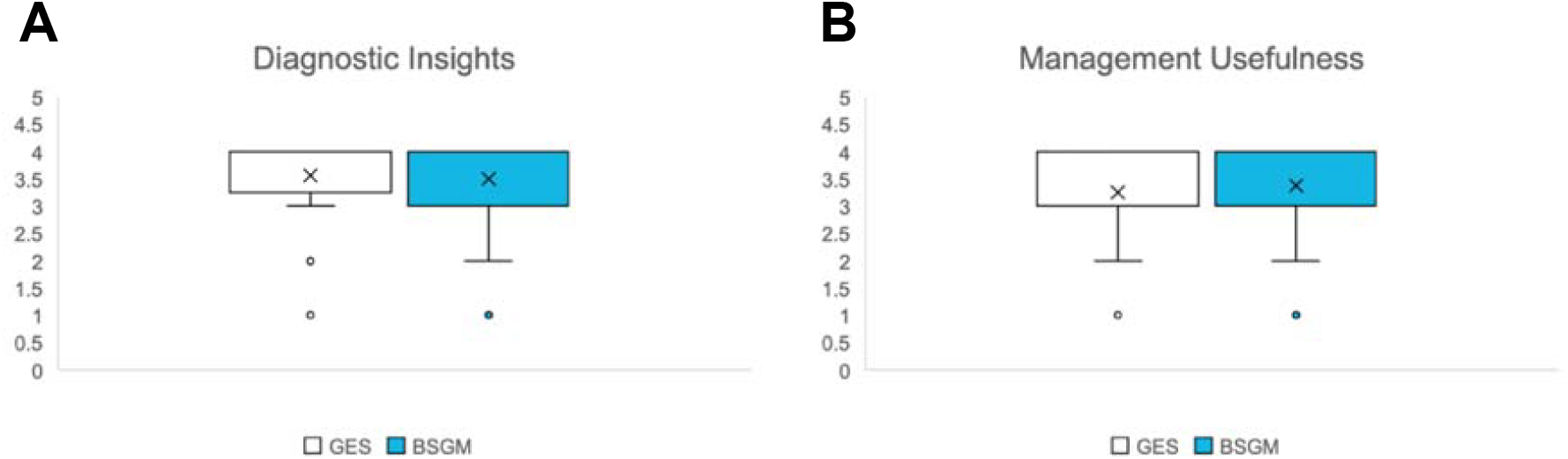
Physician-perceived clinical utility of each test result for overall diagnostic insights(A) and management usefulness (B). Statistical comparisons were analyzed using paired t test.

## Discussion

This study presents a prospective, randomized investigation of how GES and BSGM test results influence clinical management decisions in patients with suspected gastroparesis. Our findings show that combined GES+BSGM testing significantly enhanced diagnostic precision and management confidence compared to either test alone. Prior to testing, >80% of patients received multiple potential diagnoses based solely on patient history. After review of both test results, clinicians were able to establish a single definitive diagnosis in 50% of cases and >60% of patients received additional changes in management decisions, which substantially increased confidence in both diagnosis and management.

The complementary nature of these two testing modalities is a key finding. GES and BSGM detect different physiological abnormalities with minimal overlap, consistent with their assessment of distinct aspects of gastric function ^19^. While GES primarily measures the emptying rate of stomach contents, which is a higher order process ^40,41^, BSGM evaluates gastric myoelectrical activity and symptom patterns ^12,15^. Interestingly, despite measuring different physiological parameters, clinicians perceived alignment between test results in 81% of cases, leading to consistent clinical decision-making. This suggests that although the tests identify different abnormalities, they often point toward complementary clinical interpretations that reinforce their diagnosis and treatment plan.

While GES is the current reference standard for diagnosing gastroparesis, there are growing concerns regarding its clinical utility for guiding management decisions ^37,42^. Previous studies have shown that GES results often correlate poorly with symptoms, demonstrating lability over time and providing limited guidance in management ^7,9^. Our findings suggest that BSGM provides valuable additional information that complements GES, particularly through spectral analysis of gastric myoelectrical activity and structured symptom profiling. This is also consistent with our previous work showing that BSGM detects abnormalities in gastric function more frequently than GES ^19^.

A notable strength of our study design is that it simulates real-world clinical decision-making by having specialists evaluate test results sequentially, mirroring the typical process of diagnostic refinement in practice. The significant increase in both diagnostic and management certainty with combined testing, regardless of which test was reviewed first, underscores the value of this complementary approach.

Although subgroup analyses were limited by sample size, several patterns in management decisions aligned with previous research. Prokinetics and gastroparesis diets were typically recommended for patients with delayed emptying, as in previous studies ^25,43^, and consistent with current clinical guidelines ^1^. Neuromodulators were more commonly suggested for patients with normal emptying but continuous symptom profiles on BSGM, suggesting a potential gut-brain axis component ^12,15,26^. Iberogast was recommended in the case of meal-responsive or sensorimotor symptom profiles due to its ability to provide relief of symptoms associated with irritable bowel syndrome or functional dyspepsia ^44,45^. These observations, while preliminary, suggest that BSGM may help identify pathophysiological subgroups that could guide more targeted therapy.

The clinician-perceived clinical utility of GES and BSGM is a novel aspect of this study. Irrespective of the order, clinicians rated BSGM and GES as equally contributory to diagnostic insights and management decisions (p>0.05). Given that both measures offer valuable diagnostic information, the non-invasive nature of BSGM, coupled with the absence of radiation exposure (unlike GES), suggests that BSGM should be prioritized before GES in the diagnostic workup. This approach is further supported by previous health economic decision tree analyses, which suggests that incorporating BSGM early in the diagnostic pathway leads to reduced healthcare costs by streamlining the diagnostic process and decreasing the need for further invasive investigations ^22^. Moreover, the introduction of BSGM in this study significantly increased clinician certainty, showing that the additional information provided by BSGM is useful for guiding clinical management.

We note some limitations in the study. The sample size was relatively small, which limited in-depth subgroup analyses and may affect larger generalizability. Secondly, this was a single-center study with a limited number of participating clinicians, potentially introducing center-specific bias in management approaches. Despite these limitations, this study demonstrates the real-life impact of combined BSGM and GES test results in diagnosing and treating patients, reflecting management decision-making in tertiary practices, whilst also providing valuable preliminary data to guide larger, multi-center investigations. Thirdly, whilst similar clinician-perceived clinical utility questionnaires were used in previous studies ^37,38^, these questionnaires have not been formally validated in the current patient cohort, and therefore, are subject to bias. Another limitation is that while we assessed changes in clinical management decisions, we did not evaluate whether these changes led to improved patient outcomes and decreases in healthcare utilization. This is an ongoing area of evaluation.

## Conclusion

Combined GES+BSGM profiling influenced clinical management decisions, significantly enhancing diagnostic precision and management certainty in patients with suspected gastroparesis. These complementary tests support refined diagnoses and phenotype-tailored interventions. The findings illustrate the value of integrating BSGM alongside GES in the clinical evaluation of patients with complex gastroduodenal symptoms.

## Data Availability

Data used for analysis will be made available on reasonable request, conditional on ethical approvals.

