## Supplementary figures and images for "Combined Gastric Alimetry® and gastric emptying scintigraphy testing increases clinician certainty in the diagnosis and management of suspected gastroparesis"

### Supplementary Figure 1

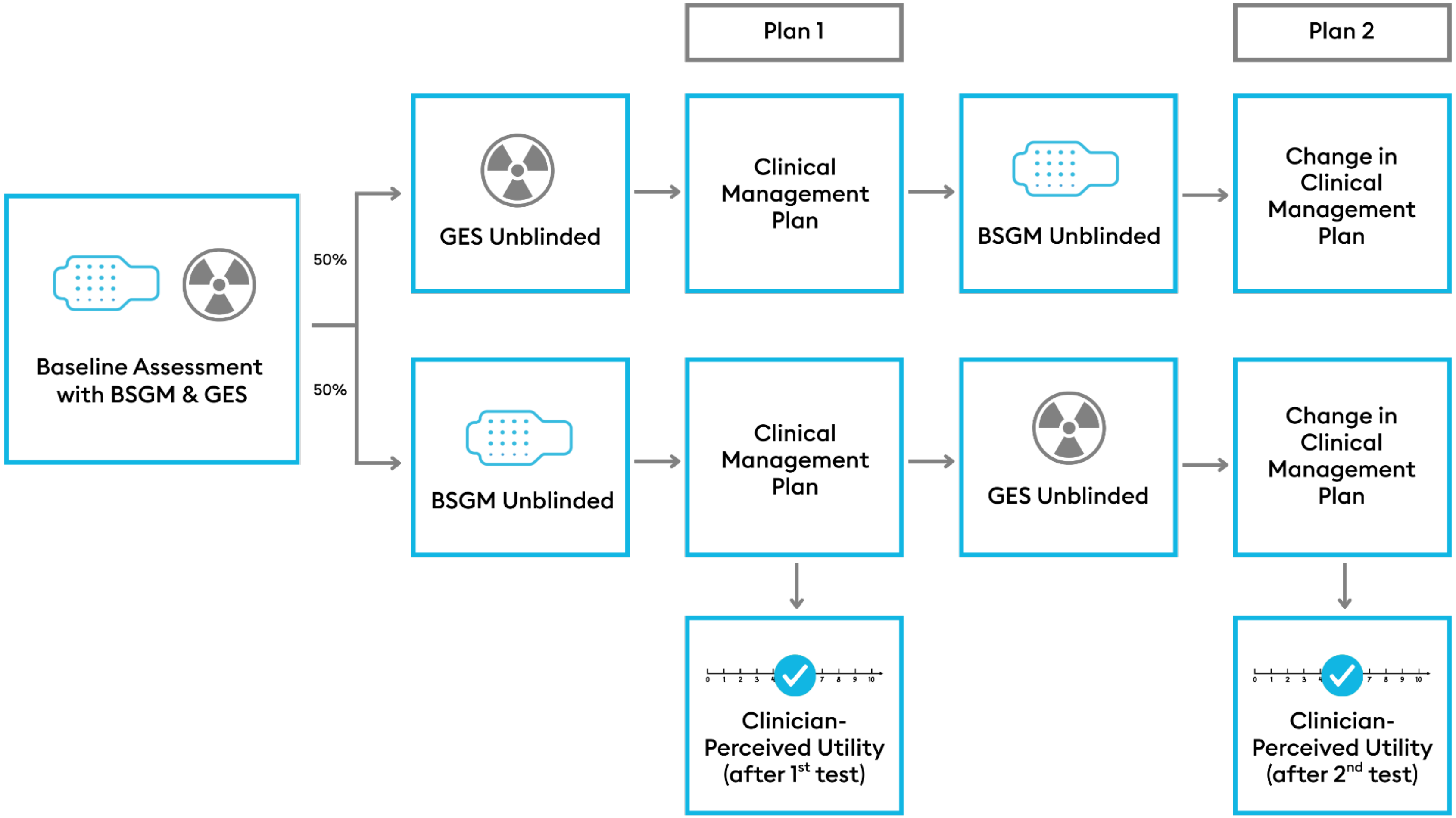


Supplementary Figure 1: Flow chart detailing the study design.
