## Supplementary Table 1 for "Combined Gastric Alimetry® and gastric emptying scintigraphy testing increases clinician certainty in the diagnosis and management of suspected gastroparesis"

Supplementary Table 1: Diagnosis and management changes for patient cohort. GES=gastric emptying scintigraphy; BSGM=body surface gastric mapping; FD=Functional Dyspepsia; GP=Gastroparesis; CNVS=chronic nausea and vomiting syndrome; CHS= Cannabinoid hyperemesis syndrome; CVS=Cyclic vomiting syndrome; ARFID=Avoidant/restrictive food intake disorder; IBS= Irritable bowel syndrome; MALS= Median Arcuate Ligament Syndrome.

| **GES then BSGM** | | | | | | | | | | | | | | |
|---|---|---|---|---|---|---|---|---|---|---|---|---|---|---|
| **Gender** | **Age Range** | **BMI** | **GES Status** | **BSGM Spectral Phenotype** | **BSGM Symptom Phenotype** | **Alignment between test results (yes/no)** | **If no: which test result best reflected the clinical diagnosis / management plan?** | **Baseline Diagnosis** | **Change in diagnosis after first unblinding** | **Change in management after first unblinding** | **Change in diagnosis after second unblinding** | **Change in management after second unblinding** | **Final Diagnosis** | **Final Plan** |
| F | 56-60 | 51.8 | Delayed | Normal | Continuous | no | GES | FD/Drug-induced/diabetic GP | No | Stop Semaglutide and refer to endocrinologist + Metoclopramide 10 mg PRN + GP diet | Yes (added FD) | No change | FD + GP | Stop Semaglutide and refer to endocrinologist + Metoclopramide + GP diet |
| F | 56-60 | 23.3 | Normal | Delayed Meal Response Ratio | Mixed (Meal-responsive + Continuous) | yes | | Suspected/diabetic GP | Yes (added CNVS and dysmotility) | Ondansetron PRN | Yes (changed from CNVS to FD) | Add Amitriptyline + Iberogast | FD + Gastric Dysmotility | Ondansetron + Amitriptyline + Iberogast |
| F | 71-75 | 28.2 | Rapid | Normal | Continuous | yes | | Suspected GP/acute illness response | Yes (added dumping syndrome) | Dumping syndrome diet + refer to dietician | No | Remove dumping syndrome diet + refer to geriatrician | Acute illness response | Refer to dietician + refer to geriatrician |
| F | 26-30 | 21.6 | Normal | Low-Amplitude | Meal-responsive | yes | | CNVS/Suspected GP/slow transit constipation | No | Iberogast + Ondansetron + Movicol + diet recommendation for small frequent meals | Yes (added FD) | No change | FD + CNVS + Slow Transit Constipation | Iberogast + Ondansetron + Movicol + diet recommendation for small frequent meals |
| F | 31-35 | 26.8 | Normal | Normal | Meal-responsive | yes | | FD/suspected GP | No | Reinforce low FODMAP diet + | No | Add referral to manometry/24 | FD | Reinforce low FODMAP diet + |

| | | | | | | | | | | | | | | |
|---|---|---|---|---|---|---|---|---|---|---|---|---|---|---|
| | | | | | | | [redacted] | | | refer to dietician | | hr pH study | | refer to dietician + referral to manometry/24 hr pH study |
| F | 21-25 | 21.2 | Normal | Normal | Continuous | yes | [redacted] | FD/suspected GP/ARFID | No | Amitriptyline 10 mg TDS + Iberogast TDS + refer to psychologist | No | Add Esomeprazole 40 mg OD | FD+ARFID | Amitriptyline + Iberogast + refer to psychologist + Esomeprazole |
| F | 66-70 | 26 | Normal | Normal | Mixed (Sensorimotor + Continuous + Post-Gastric) | yes | [redacted] | FD/Suspected GP/Slow transit constipation/IBS | No | Ondansetron 4 mg PRN + Coloxyl 120 mg PRN + low FODMAP diet | No | Add Iberogast TDS + refer to psychologist | FD + Slow Transit Constipation | Ondansetron + Coloxyl + low FODMAP diet + Iberogast + refer to psychologist |
| M | 36-40 | 18.9 | Normal | Delayed Meal Response Ratio | No symptoms on day | no | BSGM | CNVS/CHS/Suspected GP/alcohol addiction | No | Metoclopramide 10 mg PRN + advice for ongoing substance abstinence + re-attempt gastroscopy | No | Add refer to psychologist | CNVS + CHS + Alcohol Addiction | Metoclopramide + advice for ongoing substance abstinence + re-attempt gastroscopy + refer to psychologist |
| **BSGM then GES** | | | | | | | | | | | | | | |
| F | 46-50 | 31.2 | Normal | Normal | Post-gastric | yes | [redacted] | CNVS/Suspected GP | Yes (added FD) | Metoclopramide 10 mg PRN + low FODMAP diet | No | No change | FD | Metoclopramide + low FODMAP diet |
| M | 18-20 | 27.6 | Normal | High Frequency | Meal-responsive | yes | [redacted] | CVS/Suspected GP | No | Amitriptyline + Ondansetron + Refer to dietician | No | No change | CVS | Amitriptyline + Ondansetron + Refer to dietician |
| F | 18-20 | 17 | Normal | Low GA-RI | Sensorimotor | yes | [redacted] | FD/Suspected GP | No | Ondansetron + refer to dietician | Yes (added Gastric Dysmotility) | Add Iberogast | FD + Gastric Dysmotility | Ondansetron + refer to dietician + Iberogast |
| F | 26-30 | 15.6 | Normal | High-Amplitude | Mixed (Sensorimotor + Continuous) | no | GES | Suspected GP | No | Metoclopramide 10 mg TDS + Pantoprazole 40 mg OD | Yes (changed from GP to FD) | Add reinforce low FODMAP diet | FD | Metoclopramide + Pantoprazole + reinforce low FODMAP diet |
| M | 18-20 | 21.3 | Delayed | Low GA-RI | No symptoms on day | yes | [redacted] | FD/Suspected GP | No | Ondansetron 4 mg PRN | No | Add Domperidone | GP | Ondansetron + Domperidone + |

| | | | | | | | | | | | | | | |
|---|---|---|---|---|---|---|---|---|---|---|---|---|---|---|
| | | | | | | | [redacted] | | | | | + GP diet + refer to dietician | | GP diet + referral to dietician |
| F | 18-20 | 16.4 | Normal | Low GA-RI | No symptoms on day | yes | [redacted] | FD/Suspected GP | No | Ondansetron 4-8 mg PRN + refer to dietician | No | Add refer to psychologist + gut hypnotherapy | FD | Ondansetron + refer to dietician + refer to psychologist + gut hypnotherapy |
| F | 61-65 | 30 | Delayed | Normal | Mixed (Meal-responsive + Continuous) | yes | [redacted] | FD/Suspected GP | No | Gut hypnotherapy + probiotics | No | Add Domperidone 10 mg TDS | FD+GP | Gut hypnotherapy + probiotics + Domperidone |
| F | 26-30 | 14.5 | Normal | Normal | Continuous | yes | [redacted] | Suspected GP/MALS | No | Refer to upper GI surgeon for recurrent MALS treatment | No | No change | MALS | Refer to upper GI surgeon for recurrent MALS treatment |
